# A Home-Based Nurturing Care Intervention for Children born HIV-Exposed but Uninfected in Zambia: A Randomized Clinical Trial

**DOI:** 10.64898/2026.08.10.26360068

**Authors:** Ethan M. Zulu, Peter C. Rockers, Leah Forman, Namwiya Musonda, Charles Nthele, Frazer Shimaingwa, Megan Bartrum, Carol B. Masempela, Donald M. Thea, Julie M. Herlihy

## Abstract

**Introduction:** Research demonstrates neurodevelopmental differences in children who are HIV-exposed, but uninfected (CHEU) compared to their peers born unexposed. CHEUs are at higher risk for prematurity and low birth weight, both known risk factors for developmental delays. Despite increasing evidence, HIV exposure remains underrecognised as a risk factor for poor development by policy-makers and early childhood development (ECD) programs. We trialed an ECD intervention for CHEUs based on the Nurturing Care Framework developed by WHO, UNICEF, and the World Bank to improve neurodevelopmental outcomes in this vulnerable population.

**Design:** We conducted a randomized controlled trial of an ECD intervention for mothers living with HIV and their children born HIV-exposed, aged six to 24 months, living in peri-urban Lusaka. The intervention was delivered through bi-weekly home visits over 18 months using the curriculum adapted from the Nurturing Care Framework. The curriculum addressed nutrition, maternal wellness, child health, developmental milestones, and interactive caregiving.

**Method:** The primary outcome was measured by the Malawi Developmental Assessment Tool (MDAT) at six and 18 months post enrollment, which assesses developmental scores via a context-validated, direct observation tool across four domains: gross motor, fine motor, language, and social development. Participants were also assessed at six, 12 and 18 months post enrollment using the Caregiver Reported Early Development Instrument (CREDI). MDAT assessors were blinded to the intervention and control arm.

**Results:** We enrolled 308 mother-infant pairs, randomized 1:1 into an ECD intervention (CHEU control, n = 155; CHEU intervention, n = 153). At six months post enrollment, the intervention was associated with modest improvements in two developmental domains, particularly cognitive (Adjusted 0.28, 95% CI: 0.09 to 0.46; p= 0.004) and social-emotional development (Adjusted 0.29, 95% CI: 0.08 to 0.49; p= 0.07). At 18 month post enrollment, adjusted models demonstrated a moderate positive effect with gross motor development, with participants showing a 0.24 standard deviation higher score compared with the control group (95% CI: 0.02–0.46; p=0.034).

**Conclusion:** The findings indicate that the intervention may have a moderate impact on cognitive and social-emotional outcomes after six months, as well as on gross motor functions after eighteen months of intervention.

**Trial registration:** Clinicaltrials.gov NCT05119959

## Background

Recent estimates from the Global Burden of Disease Project state that 52.9 million (95% [UI] 48.7–57.3 million) children younger than 5 years have developmental disabilities, with 95% residing in low- and middle-income countries (LMIC) [1,2]. Sub-Saharan Africa has a disproportionate number of children with neurodevelopmental vulnerability, including children who are HIV-exposed but uninfected (CHEU) [3]. Evidence comparing neurodevelopment for CHEUs to peers who were non-exposed is conflicting. A study from South Africa found increased odds of cognitive and motor delay at twelve months of age [4]. Similarly, a study from Botswana showed expressive language delay at two years of age. In contrast, studies in Uganda, Malawi, and Zambia did not find any difference in neurodevelopmental delay between CHEU compared to children who were HIV unexposed (CHU) [5,6]. Differences in evaluation tools, maternal HIV disease stage, antiretroviral (ARV) regimens, ARV exposure duration, geographical location, and socio-economic context may account for the observed variability in results [6].

Early childhood development (ECD) interventions that focus on responsive parenting, caregiver well-being, and cognitive stimulation may improve developmental scores when delivered with high fidelity and on platforms accepted by the community [7]. The Nurturing Care Framework developed by World Health Organisation (<u>WHO</u>), United Nation International Children’s Emergency Fund (UNICEF) and the World Bank, in collaboration with the Partnership for Maternal, Newborn & Child Health and the Early Childhood Development Action Network, provides an integrative framework addressing five components: maternal and parental well-being, nutrition counseling on young infant and child feeding, early brain stimulation, responsive caregiving and ensuring safe environment for infants [8].

The Ministry of Health in Zambia has launched ECD initiatives, including in-service training packages and a mass-media playful parenting campaign titled “I play, I learn, I thrive” [9]. However, HIV exposure is not recognized as a risk factor for developmental vulnerability. Given the demonstrated success of community-based ECD interventions among other vulnerable populations, such as premature, stunted, or malnourished children, we hypothesized that this intervention would also be effective for CHEU to support healthy development. This study aimed to evaluate the impact of a bi-weekly ECD program, which delivered a locally adapted curriculum based on UNICEF’s Nurturing Care Framework, on neurodevelopmental outcomes among children who are HIV-exposed but uninfected (CHEU).

## Method

### Setting

The study was conducted at Chawama First Level Hospital in Lusaka between 1^st^ November 2021 and 31^st^ October 2024. This facility serves as the primary health care provider for Chawama, one of the largest peri-urban townships in Lusaka. Option B+ prevention of vertical transmission services are integrated into routine antenatal care (ANC), and women who test positive for HIV are offered immediate combination antiretroviral therapy (cART) within the same hospital. The hospital delivers approximately 8,000 babies annually, with an estimated 17% of these infants born to mothers living with HIV (approximately 1,360 per year).

### Participants

Eligibility criteria included enrollment in the Zambia Infant Cohort Study (ZICS), age of at least 18 years, and either being pregnant or having an infant younger than nine months as of February 2022. Eligible participants were women living with HIV, and their infants were classified as children born HIV-exposed but uninfected (CHEU). The mother-infant dyad’s HIV status was determined using DNA polymerase chain reaction (DNA-PCR) testing conducted within the parent study (ZICS).

### Recruitment and randomisation

Study information about the Zambia Infant Cohort Study-Optimising Brains for Surviving and Thriving (ZICS-BOOST) was provided to families who had graduated from the Zambia Infant Cohort Study (ZICS), a large observational cohort of 1500 pregnant women living with and without HIV enrolled in early pregnancy and followed to 6 months post-partum. Birth outcomes and infant infectious morbidity and mortality were published elsewhere [10]. Mothers with a child aged six to 24 months were invited to participate in ZICS-BOOST and underwent informed consent (FWA00000301 and FWA00000338). Randomisation of participants was computer generated by an independent study statistician. Good Clinical Practice (GCP) trained research nurses who enrolled the mother-infant dyads had no access to the random allocation sequence. The MDAT assessors were blinded to intervention and control group.

### Intervention

The intervention group participated in a bi-weekly ECD curriculum for 18 months. The curriculum was delivered over 36 visits by a Community Health Worker (CHW) trained in the Nurturing Care Framework developed by UNICEF, WHO and World Bank. Training was conducted over 2 weeks of 8 hours/day and reviewed age-tailored lessons from six to 48 months old. Other topics covered included maternal mental health, gender-based violence, pediatric nutrition and responsive caregiving. Community Health Workers conducted home visits every two weeks with caregivers and children in the intervention group. For participants who were uncomfortable receiving visits at home, the intervention was administered at the health facility in a private room. Each intervention session was scheduled to last approximately an hour and followed a structured format. Sessions were delivered using a flip chart manual, and included check-in, review of the previous lesson and the preceding two weeks, introduction of a new concept from the ZICS-BOOST curriculum, and guided demonstration of responsive caregiving, play, and caregiver–child interaction, with real-time feedback from the CHW. The curriculum addressed five core domains: good health (encompassing both physical and emotional health of caregiver and child), responsive caregiving, safety and security, optimal nutrition, and opportunities for early learning. During visits, CHWs modeled developmentally appropriate engagement strategies, discussed healthy and affordable nutrition options, and assessed the family’s overall safety and well-being. Some participants received selected sessions at health facilities instead of home visits due to stigma concerns, while intervention content, frequency, and duration remained unchanged. To further understand caregiver experiences and perceptions of the home-based care intervention, a follow-on qualitative study was conducted among participants. The qualitative findings will be presented in a separate manuscript and are not included in the analyses reported here.

CHWs did not provide clinical care or medical advice. Instead, they referred any health-related concerns to clinically trained staff at Chawama First Level Hospital. Neurodevelopmental assessments were conducted every six months by a Zambian child psychologist trained in MDAT administration.

### Control group

The control group, composed of CHEU participants, was randomized to receive the standard of care provided by the Ministry of Health (MoH). In Zambia, children were routinely monitored at Government of the Republic of Zambia (GRZ) clinics for growth and immunizations in accordance with the MoH-approved schedule. Under the MoH standard of care at the time of the study, there was no formalized routine assessment of neurodevelopment. If parents or healthcare providers identified specific developmental concerns, children were referred for specialized evaluation at Beit-CURE Hospital or at the University Teaching Hospital (UTH), Department of Pediatrics. For concerns related specifically to language or hearing, children were referred to the UTH Center of Excellence for Speech and Hearing for further assessment.

Children identified as demonstrating growth faltering were referred to the UTH Department of Pediatrics Malnutrition Clinic for comprehensive evaluation and management.

### Sample size and study design

Sample size calculations were informed by effect size estimates derived from a meta-analysis of 18 trials evaluating comparable early childhood development (ECD) interventions in general populations, which reported standardized mean differences of 0.42 SD and 0.47 SD for cognitive and language outcomes, respectively [11]. For the present study, a conservative standardized effect size (Cohen’s *d*) of 0.35 was assumed for each Malawi Developmental Assessment Tool (MDAT) domain z-score.

Under a two-arm parallel design, a total sample size of 258 participants (129 per arm) was estimated to provide 80% statistical power to detect this effect size using a two-sided test at α = 0.05. To account for an anticipated attrition rate of 12%, the sample size was conservatively inflated to 145 participants per arm, yielding a total sample size of 290 participants.

### Measurement tools

#### Malawi Development Assessment Tool (MDAT)

The MDAT is a context-validated developmental assessment tool developed for children aged 0 to 5 years in low- and middle-income countries (LMICs). It assesses four developmental domains: gross motor, fine motor, language, and social development, through direct observation and caregiver report, providing a reliable measure of early childhood development in research and clinical settings [12]. Domain scores are normed per population. MDATs were performed at six-month post enrollment and 18-month post enrollment. We compared the MDAT outcomes between the intervention and control groups using a difference of means to define the impact of the intervention. Assessments were conducted in designated quiet rooms by two trained assessors in the preferred language of the mother or caregiver, utilizing both combined direct child observation and caregiver reporting. Once a child failed six consecutive items in a domain, the assessors proceeded to the next domain until all were completed. The assessors were blinded to the intervention and control groups.

#### Caregiver Reported Early Development Instrument (CREDI)

Children were assessed using the long form of the CREDI at baseline, six, 12 and 18 months of follow-up. The CREDI is a caregiver-report instrument that evaluates observed age-specific tasks for children aged 0 to 36 months [13]. Trained researchers administered the assessment during the specified study visits. The long form consists of 109 questions, arranged in order of increasing difficulty. The number of responses required depends on the child’s age and developmental stage; each age group has a designated starting point, and the interview proceeds until five consecutive “no” answers are recorded. Scores were calculated for each subcategory (motor, cognitive, language, and socioemotional) as well as for overall development.

#### Self-reported questionnaire-20 (SRQ-20)

The SRQ-20 is a validated instrument to assess the mental health and well-being of caregivers. It is the tool of choice supported by the WHO and translated and validated in many LMIC countries, including Zambia [14]. It was administered by our research nurses at baseline, six, 12 and 18-month follow-ups. Other secondary endpoints included dietary diversity z-scores and caregiver–child interaction scores. Dietary diversity was assessed using a 24-hour caregiver recall based on the WHO and UNICEF Infant and Young Child Feeding (IYCF) guidelines [15], while caregiver–child interactions were measured using the six-item Multiple Indicator Cluster Survey (MICS) module [16].

#### Eye tracking

Eye-tracking procedures offer valuable insights into the development of cognitive, social, and emotional processing during infancy [17,18]. In this study, eye-tracking assessments were conducted in a quiet, dimly lit room at 18-month follow-up. Children were seated on their mother’s lap approximately 60 cm from a 22-inch monitor equipped with a Tobii Pro Fusion eye tracker (Tobii AB, Stockholm, Sweden). Three early cognitive function tests were administered: the visual search task, which assessed the ability to locate some culturally-appropriate target presented alone (one-object condition), among identical distractors (distractor condition), or among two types of distractors (conjunction condition); the switch-task, which evaluated the ability to learn and anticipate the side where a target would appear during preswitch and postswitch phases; and the disengagement task, which measured the ability to disengage attention from a non-face pattern or a happy or fearful face to a salient lateral stimulus. The assessor was blinded to the intervention status of the children.

#### Child Average Growth Assessment

Height was obtained using a portable stadiometer and weight using an electronic scale at baseline, six and 18 months of follow-up. Length was measured in recumbent position for children younger than 2 years, while standing height was measured for older children. Anthropometrics were obtained by experienced research nurses who received a refresher training. Children were weighed wearing light clothing. Z-scores for height-for-age, length-for-age, weight-for-age and weight-for-length were calculated using the 2006 WHO child growth standards.

### Quality Assurance

Standardized developmental assessments were conducted by a child psychologist (N.M) and an experienced trained examiner (N.K), both blinded to participants’ HIV exposure status. Assessments of children were done in quiet clinical rooms prepared for this purpose. The child psychologist conducted 39.1% of the MDAT assessment, while the trained examiner conducted 61.9%. All the assessments done by the trained examiner were reviewed by the child psychologist. MDAT forms were reviewed daily by the study team and weekly by the study statistician. During data quality checks, the study statistician identified invalid scores, which were subsequently reviewed and corrected by the two assessors. The identified discrepancies were infrequent and limited to clerical errors, including incorrect domain score summations or data-entry mistakes. These were verified against the original MDAT forms and corrected accordingly. Weekly meetings were held to address any concerns arising from the assessment procedures.

### Statistical analysis

We estimated the impact of the intervention on the primary outcomes of interest using an intention-to-treat approach. We fit a set of linear regression models to estimate unadjusted and adjusted treatment effects. Adjusted models included a set of demographic variables measured prior to the start of the intervention: child date of birth, age at study enrollment, sex, gestational age and fetal growth status, Apgar score, breastfeeding status at 20 weeks, baseline HAZ, and baseline CREDI overall z-score; mother’s age, education, ART status at child’s conception, and blood pressure status during pregnancy; household size and wealth. For all analyses, data from participants not assessed at endline were assumed to be missing completely at random and a complete-case approach was used.

### Ethics and consent

The study protocol, including the participant compensation procedures, was reviewed and approved by the University of Zambia Biomedical Research Ethics Committee (FWA00000338) and the Boston University Institutional Review Board (FWA00000301). Written informed consent was obtained from all parents/guardians before enrolment. Participants continued to receive the standard travel reimbursement for scheduled study visits throughout the follow-up period. Participants randomized to the intervention arm also received an inconvenience stipend for the 36 home visits to acknowledge the time commitment required from caregivers. These visits required caregivers to dedicate time that could otherwise be used for income-generating activities. In addition, upon completion of the study, participants received a modest retention package consisting of household items (e.g., fabric, soap, and cooking oil) valued at approximately US$5 as a token of appreciation for their continued participation. This study was registered on ClinicalTrials.gov under the code: https://clinicaltrials.gov/study/NCT05119959. Registered on November 3, 2021.

## Results

Between March 2022 and October 2024, a total of 308 mother-infant pairs were enrolled in the study (CHEU control, n = 155; CHEU intervention, n = 153). Follow-up was available for 293 (94.8%) at midline 1 (CHEU control, n = 144; CHEU intervention, n = 149), excluding a total of 16 infants who either missed the assessment or were lost-to-follow up. Further follow-up at midline 2 (CREDI assessment) was available for 264 infants (85.1%) (CHEU control, n = 123; CHEU intervention, n = 141). MDAT assessments at endline were done for a total of 289 infants (93.5%) [140 CHEU control children (89.7%) and 149 CHEU intervention children (97.3%)]. Overall, 20 (6.5%) participants had missing outcomes at endline. (Figure 1)

**Fig. 1.**
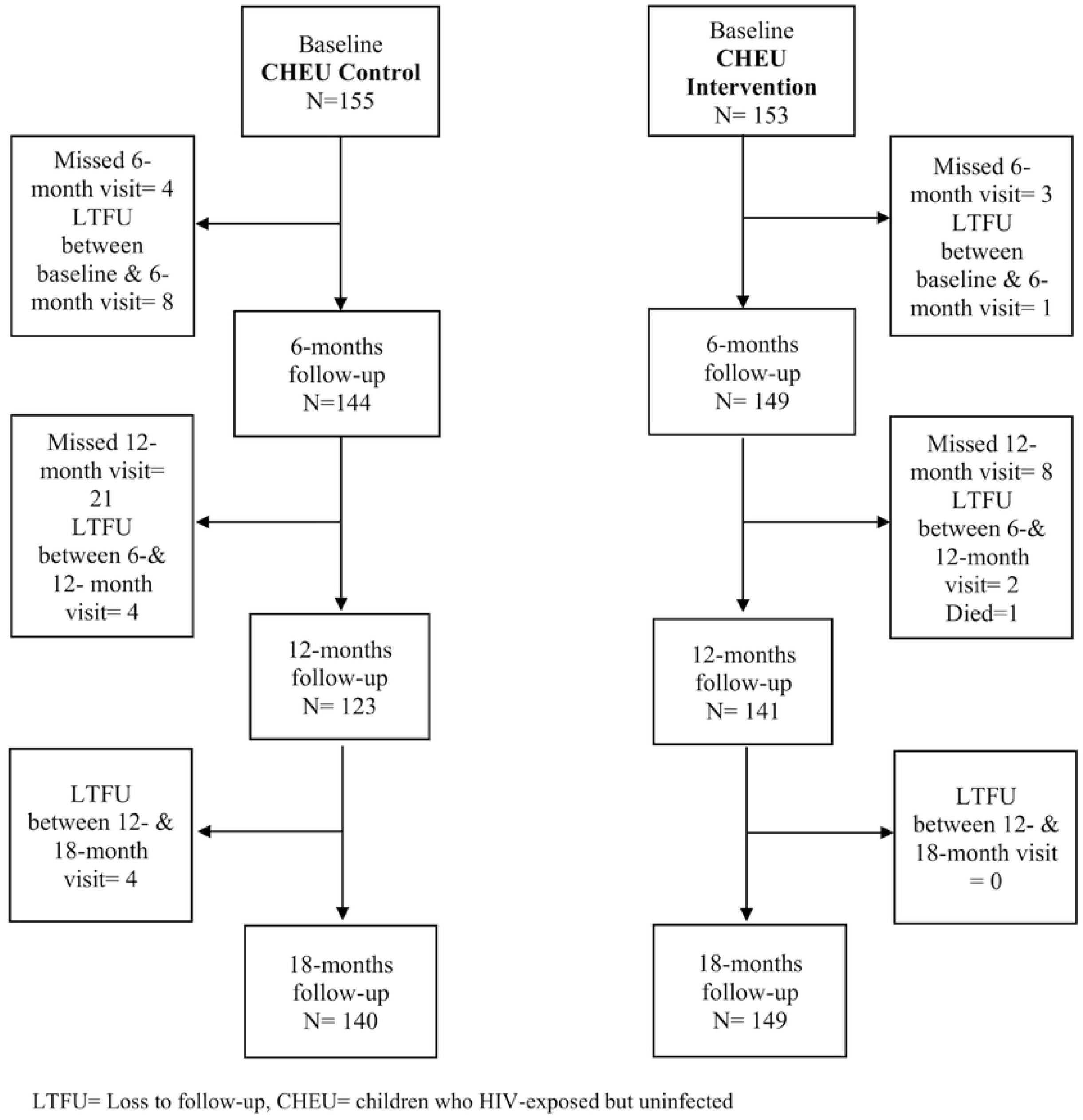
Flow diagram.

Table 1 presents the baseline characteristics of child-caregiver dyads in the study, separated into intervention and control groups. Among the 308 dyads analyzed, 153 children were assigned to the intervention group and 155 to the control group. The two groups were similar across all measured demographic variables at baseline, except that the intervention group included a slightly higher proportion of mothers in the fifth quintile compared to the control group.

**Table 1.** Maternal and children baseline characteristics.

|  |  |  | Study Group |  |
| --- | --- | --- | --- | --- |
| Variable | Response | Overall | Intervention | Control |
|  |  | 308 | 153 | 155 |
| *Maternal characteristics* |  |  |  |  |
| Age of mothers (in years) | N | 308 | 153 | 155 |
|  | Mean (Std Dev) | 29.8 (6.0) | 29.5 (5.7) | 30.0 (6.3) |
| Marital status N (%) | Other | 40 (13.0%) | 16 (10.5%) | 24 (15.5%) |
|  | Married | 268 (87.0%) | 137 (89.5%) | 131 (84.5%) |
| Employment status N (%) | Yes, full-time | 45 (14.6%) | 20 (13.1%) | 25 (16.1%) |
|  | Yes, part-time | 36 (11.7%) | 22 (14.4%) | 14 (9.0%) |
| Variable | Response | Overall | Intervention | Control |
|  | No | 227 (73.7%) | 111 (72.5%) | 116 (74.8%) |
| Education status N (%) | No formal education | 7 (2.3%) | 4 (2.6%) | 3 (1.9%) |
|  | Primary | 85 (27.6%) | 43 (28.1%) | 42 (27.1%) |
|  | Secondary | 204 (66.2%) | 99 (64.7%) | 105 (67.7%) |
|  | College (and beyond) | 12 (3.9%) | 7 (4.6%) | 5 (3.2%) |
| Zambia Wealth quintile N (%) | 1st quintile | 9 (3.0%) | 5 (3.3%) | 4 (2.6%) |
|  | 2nd quintile | 64 (21.2%) | 22 (14.7%) | 42 (27.6%) |
|  | 3rd quintile | 130 (43.0%) | 66 (44.0%) | 64 (42.1%) |
|  | 4th quintile | 76 (25.2%) | 39 (26.0%) | 37 (24.3%) |
|  | 5th quintile | 23 (7.6%) | 18 (12.0%) | 5 (3.3%) |
| Smoke during current pregnancy N (%) | No | 306 (99.4%) | 152 (99.3%) | 154 (99.4%) |
|  | Yes | 2 (0.6%) | 1 (0.7%) | 1 (0.6%) |
| Alcohol during current pregnancy N (%) | No | 263 (85.4%) | 131 (85.6%) | 132 (85.2%) |
|  | Yes | 45 (14.6%) | 22 (14.4%) | 23 (14.8%) |
| Undetectable viral load ($\leq 20$) | No | 145 (48.7%) | 70 (47.0%) | 75 (50.3%) |
|  | Yes | 153 (51.3%) | 79 (53.0%) | 74 (49.7%) |
| ART initiation timing N (%) | Pre-conception (ART > 6 months) | 236 (76.6%) | 115 (75.2%) | 121 (78.1%) |
|  | Post-conception (ART < 6 months) | 72 (23.4%) | 38 (24.8%) | 34 (21.9%) |
| *Children characteristics* |  |  |  |  |
| Birthweight (categorical) | > 2500g | 241 (78.2%) | 117 (76.5%) | 124 (80.0%) |
| | LBW (>1500g - $\leq 2500$g) | 66 (21.4%) | 35 (22.9%) | 31 (20.0%) |
| Variable | Response | Overall | Intervention | Control |
|  | VLBW (>1000g - <=1500g) | 1 (0.3%) | 1 (0.7%) | 0 (0.0%) |
| Sex of child | Male | 150 (48.7%) | 70 (45.8%) | 80 (51.6%) |
|  | Female | 158 (51.3%) | 83 (54.2%) | 75 (48.4%) |
| Small for gestation age: < 10th percentile | No | 221 (71.8%) | 107 (69.9%) | 114 (73.5%) |
|  | Yes | 87 (28.2%) | 46 (30.1%) | 41 (26.5%) |
| Delivery < 37 weeks - using BOE | No | 277 (89.9%) | 138 (90.2%) | 139 (89.7%) |
|  | Yes | 31 (10.1%) | 15 (9.8%) | 16 (10.3%) |
| Breastfeeding at 20 weeks | None | 20 (6.5%) | 10 (6.5%) | 10 (6.5%) |
|  | Any | 77 (25.0%) | 41 (26.8%) | 36 (23.2%) |
|  | Exclusive | 211 (68.5%) | 102 (66.7%) | 109 (70.3%) |
| Food diversity score | N | 308 | 153 | 155 |
|  | Mean (Std Dev) | 4.0 (2.1) | 4.1 (2.1) | 3.9 (2.2) |
| Length-for-age Z-score | N | 307 | 152 | 155 |
|  | Mean (Std Dev) | -0.6 (14.8) | 0.2 (20.9) | -1.4 (1.9) |
| Weight-for-age Z score | N | 308 | 153 | 155 |
|  | Mean (Std Dev) | -0.6 (1.2) | -0.6 (1.1) | -0.6 (1.2) |
| CREDI Overall Z-Score | N | 308 | 153 | 155 |
|  | Mean (Std Dec) | -0.2 (0.7) | -0.2 (0.8) | -0.2 (0.7) |

ART=Antiretroviral treatment, BOE= best obstetric estimation, LBW= low birthweight, VLBW= very low birthweight

Table 2 presents the impact of the intervention on developmental outcomes among children. For Malawi Developmental Assessment Tool (MDAT) outcomes, no effect was observed in the gross motor domain after six months. However, at 18 months, adjusted models demonstrated a moderate positive association with gross motor development, with participants showing a 0.24 standard deviation higher score compared with the control group (95% CI: 0.02–0.46; p=0.034). Although positive trends were observed at all measured intervals in language, fine motor, and social adaptive domains with MDAT, neither unadjusted nor adjusted models reached statistical significance. Additionally, there was no significant association between the intervention and saccadic reaction time after 18 months (Adjusted β = 0.41, 95% CI: −11.25 to 12.06; p = 0.945).

**Table 2.**
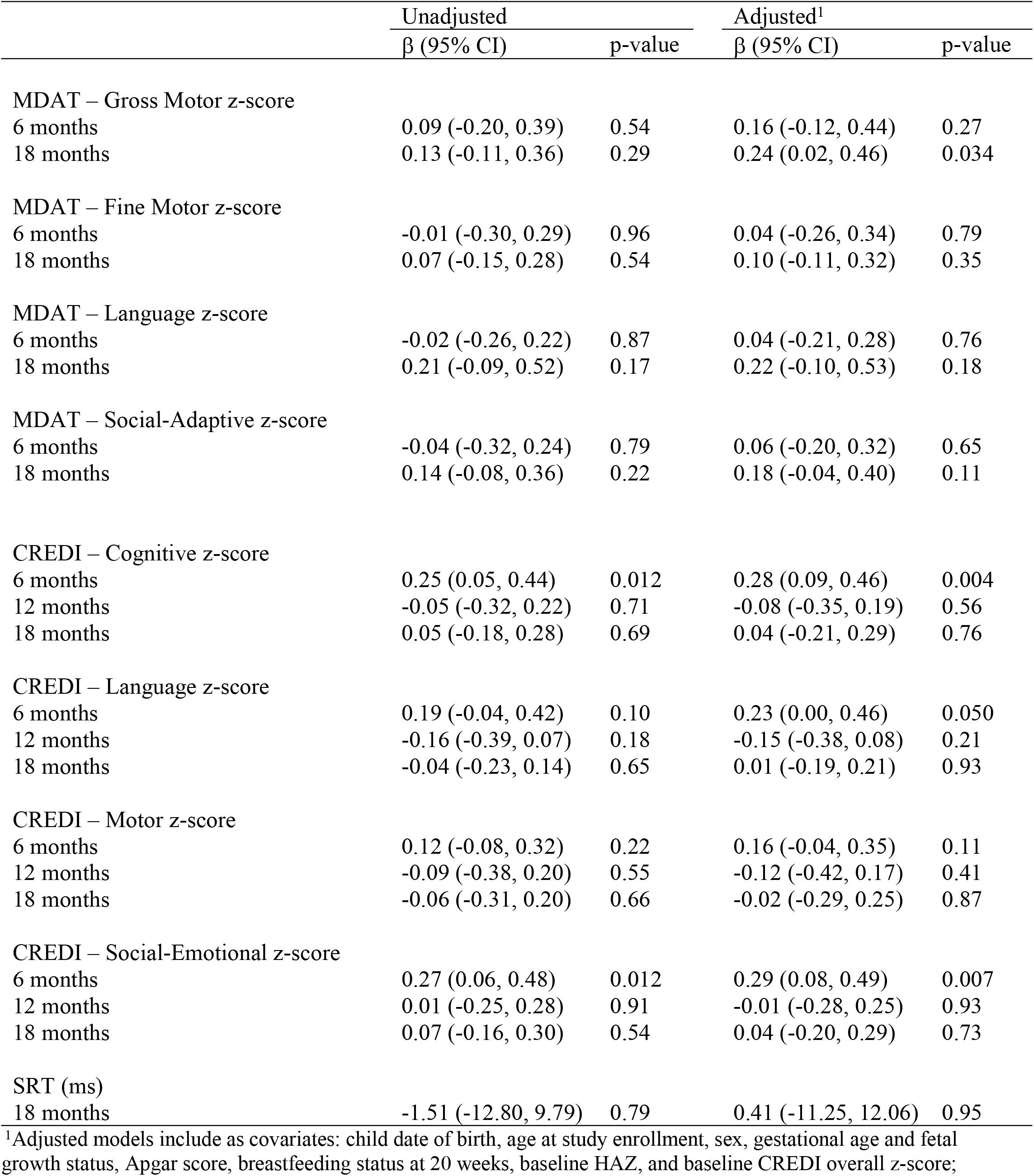
Impacts on primary outcomes.

|  | Unadjusted |  | Adjusted<sup>1</sup> |  |
| --- | --- | --- | --- | --- |
| | $\beta$ (95% CI) | p-value | $\beta$ (95% CI) | p-value |
| MDAT – Gross Motor z-score |  |  |  |  |
| 6 months | 0.09 (-0.20, 0.39) | 0.54 | 0.16 (-0.12, 0.44) | 0.27 |
| 18 months | 0.13 (-0.11, 0.36) | 0.29 | 0.24 (0.02, 0.46) | 0.034 |
| MDAT – Fine Motor z-score |  |  |  |  |
| 6 months | -0.01 (-0.30, 0.29) | 0.96 | 0.04 (-0.26, 0.34) | 0.79 |
| 18 months | 0.07 (-0.15, 0.28) | 0.54 | 0.10 (-0.11, 0.32) | 0.35 |
| MDAT – Language z-score |  |  |  |  |
| 6 months | -0.02 (-0.26, 0.22) | 0.87 | 0.04 (-0.21, 0.28) | 0.76 |
| 18 months | 0.21 (-0.09, 0.52) | 0.17 | 0.22 (-0.10, 0.53) | 0.18 |
| MDAT – Social-Adaptive z-score |  |  |  |  |
| 6 months | -0.04 (-0.32, 0.24) | 0.79 | 0.06 (-0.20, 0.32) | 0.65 |
| 18 months | 0.14 (-0.08, 0.36) | 0.22 | 0.18 (-0.04, 0.40) | 0.11 |
| CREDI – Cognitive z-score |  |  |  |  |
| 6 months | 0.25 (0.05, 0.44) | 0.012 | 0.28 (0.09, 0.46) | 0.004 |
| 12 months | -0.05 (-0.32, 0.22) | 0.71 | -0.08 (-0.35, 0.19) | 0.56 |
| 18 months | 0.05 (-0.18, 0.28) | 0.69 | 0.04 (-0.21, 0.29) | 0.76 |
| CREDI – Language z-score |  |  |  |  |
| 6 months | 0.19 (-0.04, 0.42) | 0.10 | 0.23 (0.00, 0.46) | 0.050 |
| 12 months | -0.16 (-0.39, 0.07) | 0.18 | -0.15 (-0.38, 0.08) | 0.21 |
| 18 months | -0.04 (-0.23, 0.14) | 0.65 | 0.01 (-0.19, 0.21) | 0.93 |
| CREDI – Motor z-score |  |  |  |  |
| 6 months | 0.12 (-0.08, 0.32) | 0.22 | 0.16 (-0.04, 0.35) | 0.11 |
| 12 months | -0.09 (-0.38, 0.20) | 0.55 | -0.12 (-0.42, 0.17) | 0.41 |
| 18 months | -0.06 (-0.31, 0.20) | 0.66 | -0.02 (-0.29, 0.25) | 0.87 |
| CREDI – Social-Emotional z-score |  |  |  |  |
| 6 months | 0.27 (0.06, 0.48) | 0.012 | 0.29 (0.08, 0.49) | 0.007 |
| 12 months | 0.01 (-0.25, 0.28) | 0.91 | -0.01 (-0.28, 0.25) | 0.93 |
| 18 months | 0.07 (-0.16, 0.30) | 0.54 | 0.04 (-0.20, 0.29) | 0.73 |
| SRT (ms) |  |  |  |  |
| 18 months | -1.51 (-12.80, 9.79) | 0.79 | 0.41 (-11.25, 12.06) | 0.95 |

<sup>1</sup>Adjusted models include as covariates: child date of birth, age at study enrollment, sex, gestational age and fetal growth status, Apgar score, breastfeeding status at 20 weeks, baseline HAZ, and baseline CREDI overall z-score;

After six months, the intervention was associated with modest improvements in certain developmental domains, particularly cognitive (Adjusted 0.28, 95% CI: 0.09 to 0.46; p= 0.004) and social-emotional development (Adjusted 0.29, 95% CI: 0.08 to 0.49; p= 0.07). However, these effects were generally not sustained at 12 and 18 months post-intervention. No significant effects were observed for language and motor domains in the intervention group in adjusted models using the Caregiver Reported Early Development Instruments (CREDI).

Table 3 presents the effects of the intervention on diet diversity and child interactions. Across all follow-up time points, there was no statistically significant effect of the intervention on food diversity scores. However, consistent positive effects of the intervention on caregiver-child interactions were observed at each assessment. After six months, children in the intervention group had higher MICS interaction scores compared to controls (adjusted β = 0.33, 95% CI: 0.08 to 0.57; p = 0.010). This association persisted after 12 months (adjusted β = 0.37, 95% CI: 0.08 to 0.65; p = 0.012) and remained statistically significant after 18 months (adjusted β = 0.28, 95% CI: 0.02 to 0.54; p = 0.032).

**Table 3.** Impacts on secondary outcomes.

|  | Unadjusted |  | Adjusted<sup>1</sup> |  |
| --- | --- | --- | --- | --- |
| | $\beta$ (95% CI) | p-value | $\beta$ (95% CI) | p-value |
| Diet diversity score |  |  |  |  |
| 6 months | 0.35 (-0.04, 0.74) | 0.080 | 0.33 (-0.08, 0.75) | 0.12 |
| 12 months | 0.09 (-0.29, 0.47) | 0.65 | 0.02 (-0.38, 0.42) | 0.93 |
| 18 months | 0.02 (-0.31, 0.36) | 0.89 | 0.09 (-0.26, 0.45) | 0.60 |
| Child interactions (MICS score) |  |  |  |  |
| 6 months | 0.41 (0.15, 0.66) | 0.002 | 0.33 (0.08, 0.57) | 0.010 |
| 12 months | 0.42 (0.14, 0.69) | 0.003 | 0.37 (0.08, 0.65) | 0.012 |
| 18 months | 0.22 (-0.02, 0.45) | 0.076 | 0.28 (0.02, 0.54) | 0.032 |

<sup>1</sup>Adjusted models include as covariates: child date of birth, age at study enrollment, sex, gestational age and fetal growth status, Apgar score, breastfeeding status at 20 weeks, baseline HAZ, and baseline CREDI overall z-score; mother's age, education, ART status at child's conception, and blood pressure status during pregnancy; household size and wealth.

## Discussion

This study estimated the effect of a bi-weekly ECD program, which delivered a locally adapted curriculum based on the Nurturing Care Framework, on neurodevelopment among children exposed to HIV but uninfected (CHEUs). As part of the earlier adaptation process, the 36 lesson curriculum was tailored for a one-on-one home visitation program providing support to families in child development, nutrition counseling, maternal wellness and HIV medication adherence. This work represents the first steps towards understanding the effectiveness of early childhood interventions (ECD) on a growing population of CHEUs. Zambia has been scaling up ECD interventions but CHEUs are not currently targeted. Our prior research demonstrated that CHEUs are at increased risk for prematurity and low birth weight but not at increased risk for neurodevelopment [10]. This finding was in a cohort of women with well controlled HIV disease, early ART initiation and excellent medication adherence. Our trial demonstrated limited yet potentially meaningful domain-specific developmental benefits from the intervention, particularly for cognitive and social-emotional domains during early infancy and for gross motor skills in later infancy. We demonstrated a positive, significant effect on caregiver-child interactions which is critical to long term development. These results seem consistent with other research that found an impact of caregiver-child interactions on developmental outcomes [19]. While the intervention showed a link to an increase in developmental outcomes in CHEUs, the magnitude of the effect was moderate, particularly in cognitive, social-emotional and gross motor.

The consistent positive effect of increased caregiver-child interactions seen in the intervention presents a possible mechanism through which the intervention could have influenced a moderate improvement in cognitive and social-emotional outcomes. Differences observed in certain domains, particularly language, motor, and social-emotional or social-adaptive areas, between the CREDI and MDAT assessments in this cohort likely reflect differences in assessment methodology. The CREDI is a self-reported questionnaire designed to capture broader aspects of early developmental functioning, whereas the MDAT is a directly observed assessment focused on specific developmental tasks. Therefore, discrepancies at 18 months in language, motor, and social-emotional domains between the MDAT and CREDI instruments are more indicative of differences in measurement approaches than of inconsistencies in the underlying developmental associations.

Our study had several limitations. First, CREDI assessments relied on caregiver self-report and may therefore have been subject to social desirability bias. However, this limitation does not apply to our primary outcome, which was assessed using a direct observation tool. Second, some caregivers chose to receive the intervention at the health facility rather than through home visits after initial contact with community health workers. This may have been related to concerns about stigma; however, we did not collect data on the reasons for this preference and were therefore unable to conduct subgroup analyses to assess its potential impact on intervention delivery or outcomes.

Despite these limitations, the study had notable strengths. These include the prospective cohort design with extensive data from the previous parent study, accurate gestational dating by ultrasound, precise determination of maternal combined antiretroviral treatment (cART) timing, high participant follow-up (93.5%), and sociodemographic balance between comparison groups. The blinding of MDAT outcome assessors and the use of intention-to-treat analysis further strengthen the study.

## Conclusion

This study set out to evaluate the impact of a bi-weekly community health worker home visitation program, which delivered a locally adapted curriculum based on UNICEF’s Nurturing Care Framework, on neurodevelopmental outcomes among children born HIV-exposed but uninfected (CHEUs). In summary, our findings, indicate that the intervention may have a moderate impact on cognitive and social-emotional outcomes after six months, as well as on gross motor functions after 18 months of intervention. Future studies should follow these children longitudinally to determine whether ECD interventions can affect school readiness and literacy. Larger randomized controlled trials are needed to provide more definitive evidence. Future research should also investigate more objective methods for assessing neurodevelopment, such as eye-tracking technology and evaluate the role of father engagement on neurological outcomes in children who are HIV-exposed but uninfected. CHEUs represent a major portion of the population and although our previous work showed similar neurodevelopment to their peers who are HIV-unexposed, important questions remain on regarding best to support their development. These questions are particularly urgent as programs supporting the prevention of vertical HIV transmission face potential disruptions due to declining or uncertain funding.

## Data Availability

The minimal data set and accompanying code is provided with the article

## Acknowledgements

We would like to thank all the participants who took part in the study, as their involvement was essential to this research. We also thank Whitney Morreau for proofreading.

